# No patient is the same; lessons learned from antibody repertoire profiling in hospitalized severe COVID-19 patients

**DOI:** 10.1101/2022.12.23.22283896

**Authors:** Albert Bondt, Max Hoek, Kelly Dingess, Sem Tamara, Bastiaan de Graaf, Weiwei Peng, Maurits A. den Boer, Mirjam Damen, Ceri Zwart, Arjan Barendregt, Danique M.H. van Rijswijck, Marloes Grobben, Khadija Tejjani, Jacqueline van Rijswijk, Franziska Völlmy, Joost Snijder, Francesca Fortini, Alberto Papi, Carlo Alberto Volta, Gianluca Campo, Marco Contoli, Marit J. van Gils, Savino Spadaro, Paola Rizzo, Albert J.R. Heck

**Affiliations:** Biomolecular Mass Spectrometry and Proteomics, Bijvoet Center for Biomolecular Research and Utrecht Institute for Pharmaceutical Sciences, University of Utrecht, Padualaan 8, 3584 CH, Utrecht, The Netherlands; Netherlands Proteomics Center, Padualaan 8, 3584 CH, Utrecht, The Netherlands; Department of Medical Microbiology and Infection Prevention, Amsterdam UMC, University of Amsterdam, Amsterdam Institute for Infection and Immunity, Meibergdreef 9, 1105 AZ, Amsterdam, The Netherlands; Maria Cecilia Hospital, GVM Care & Research, Cotignola, Italy; Respiratory Section, Department of Translational Medicine, University of Ferrara, Ferrara, Italy and Respiratory Disease Unit, Azienda Ospedaliero-Universitaria di Ferrara, Italy; Department of Translational Medicine University of Ferrara, Ferrara, Italy and Intensive Care Unit, Azienda Ospedaliero-Universitaria di Ferrara, Italy; Cardiology Unit, Azienda Ospedaliero-Universitaria di Ferrara, University of Ferrara, Ferrara, Italy; Department of Translational Medicine and Laboratory for Technology of Advanced Therapies (LTTA), University of Ferrara, Italy

**Keywords:** COVID-19, IgG1, IgA1, mass spectrometry, immunoglobulin repertoire, human antibodies, tocilizumab, personalized serology

## Abstract

Here, by using mass spectrometry-based methods IgG1 and IgA1 clonal repertoires were monitored quantitatively and longitudinally in more than 50 individual serum samples obtained from 17 COVID-19 patients admitted to intensive care units because of acute respiratory distress syndrome. These serological clonal profiles were used to examine how each patient reacted to a severe SARS-CoV-2 infection. All 17 donors revealed unique polyclonal repertoires and changes after infection. Substantial changes over time in the IgG1 and/or IgA1 clonal repertoires were observed in individual patients, with several new clones appearing following the infection, in a few cases leading to a few very high abundant IgG1 and/or IgA1 clones dominating the repertoire. Several of these clones were *de novo* sequenced through combinations of top-down, middle-down and bottom-up proteomics approaches. This revealed several sequence features in line with sequences deposited in the SARS-CoV-specific database of antibodies. In other patients, the serological Ig profiles revealed the treatment with tocilizumab, as after treatment, this IgG1-mAb dominated the serological IgG1 repertoire. Tocilizumab clearance could be monitored and a half-life of approximately 6 days was established in these patients. Overall, our longitudinal monitoring of IgG1 and IgA1 repertoires of individual donors reveals that antibody responses are highly personalized traits of each patient, affected by the disease and the chosen clinical treatment. The impact of these observations argues for a more personalized and longitudinal approach in patients’ diagnostics, both in serum proteomics as well as in monitoring immune responses.

## Introduction

More than 30% of the proteins in our blood are immunoglobulins. Of these, IgG1 molecules account for 20-30% and IgA1 for 5-10%. Immunoglobulins, also known as antibodies, are glycoprotein molecules produced by plasma cells and they act as a key part of the adaptive immune response by specifically recognizing and binding to antigens derived from bacteria or viruses, thereby initiating and/or aiding in their destruction. Each human can produce a vast variety of distinct antibody-producing B cell clones, with estimates ranging from 10^13^ to 10^18^ (Schroeder 2006, Briney, Inderbitzin et al. 2019). This repertoire allows humans to adequately respond to attacks by the huge variety of pathogens and other foreign elements we daily encounter. However, as shown previously by others and us, at a given moment in time there are likely only thousands of different antibodies circulating in detectable amounts in our blood, and typically the top 50 most abundant Ig clones account for up to >90% of the complete Ig repertoire (Wine, Boutz et al. 2013, Lee, Paparoditis et al. 2019, Bondt, Dingess et al. 2021, Bondt, Hoek et al. 2021). Recently, we developed methods to directly affinity-purify all IgG molecules from serum or plasma, and subsequently cleave of the IgG1-Fab fragments (that harbor the important antigen recognizing CDR regions), subjecting them to mass analysis by intact Fab profiling LC-MS (FP-MS). Each FP-MS trace contained a few hundred unique signals (based on mass and retention time) that we considered unique clones. By spiking in recombinant IgG1 mAb standards, we were able to monitor the serum concentration of each clone in the repertoire over time (Bondt, Hoek et al. 2021). Next, we adapted the FP-MS sample preparation to target specifically IgA1, using a different affinity-capture resin, another protease to produce the Fab fragments, and recombinant IgA1 mAb standards (Bondt, Dingess et al. 2021). By monitoring these IgG1 and IgA1 repertoires, we observed that human plasma IgG1 and IgA1 repertoires are relatively simple (dominated by just a few hundred different clones), but also unique and highly personalized as we rarely observed the same clones in more than one donor. Moreover, in healthy donors we found that the IgG1 and IgA1 repertoires were very stable over time. In contrast, in patients experiencing a septic episode we noticed substantial changes in the IgG1 repertoires (Bondt, Hoek et al. 2021).

In the present study, we set out to follow up on these earlier findings by focusing on a real-life small cohort of patients, all infected with the same pathogen, namely SARS-CoV-2. This cohort was not designed for this study but is a subgroup of patients from the ATTAC-Co study (registered at www.clinicaltrials.gov, number NCT04343053). The ATTAC-Co study is an investigator-initiated, prospective, single-center study recruiting patients admitted to the intensive care unit (ICU) at the University Hospital of Ferrara, Italy, because of COVID-19-associated acute respiratory distress syndrome (CARDS) at the onset of the pandemic between April and May 2020. We selected from this cohort 17 patients for which three different longitudinal blood samplings were made: just after admission to the ICU (T1), after 7 ± 3 d (T2), and after 14 ± 4 d (T3). Earlier, we sampled the serum proteomes of these patients, which allowed us to define a serum proteome signature that could be used to predict mortality in severe COVID-19 patients (Vollmy, van den Toorn et al. 2021). This serum proteome signature was confirmed by similar studies appearing at the same time (Demichev, Tober-Lau et al. 2021, Geyer, Arend et al. 2021).

Of the 17 patients whose serum we sampled longitudinally in this study 5 did not survive. At the early onset of the pandemic, *i*.*e*. when these patients were hospitalized, it was still very much unclear what would be the best treatments for COVID-19 patients and therefore it may come as no surprise that patients were not treated all in the same manner, *e*.*g*. they encountered variable numbers of plasma, red blood cell or platelet transfusions, and various forms of treatment with immune-suppressing medication (Figure 1).

**Figure 1.**
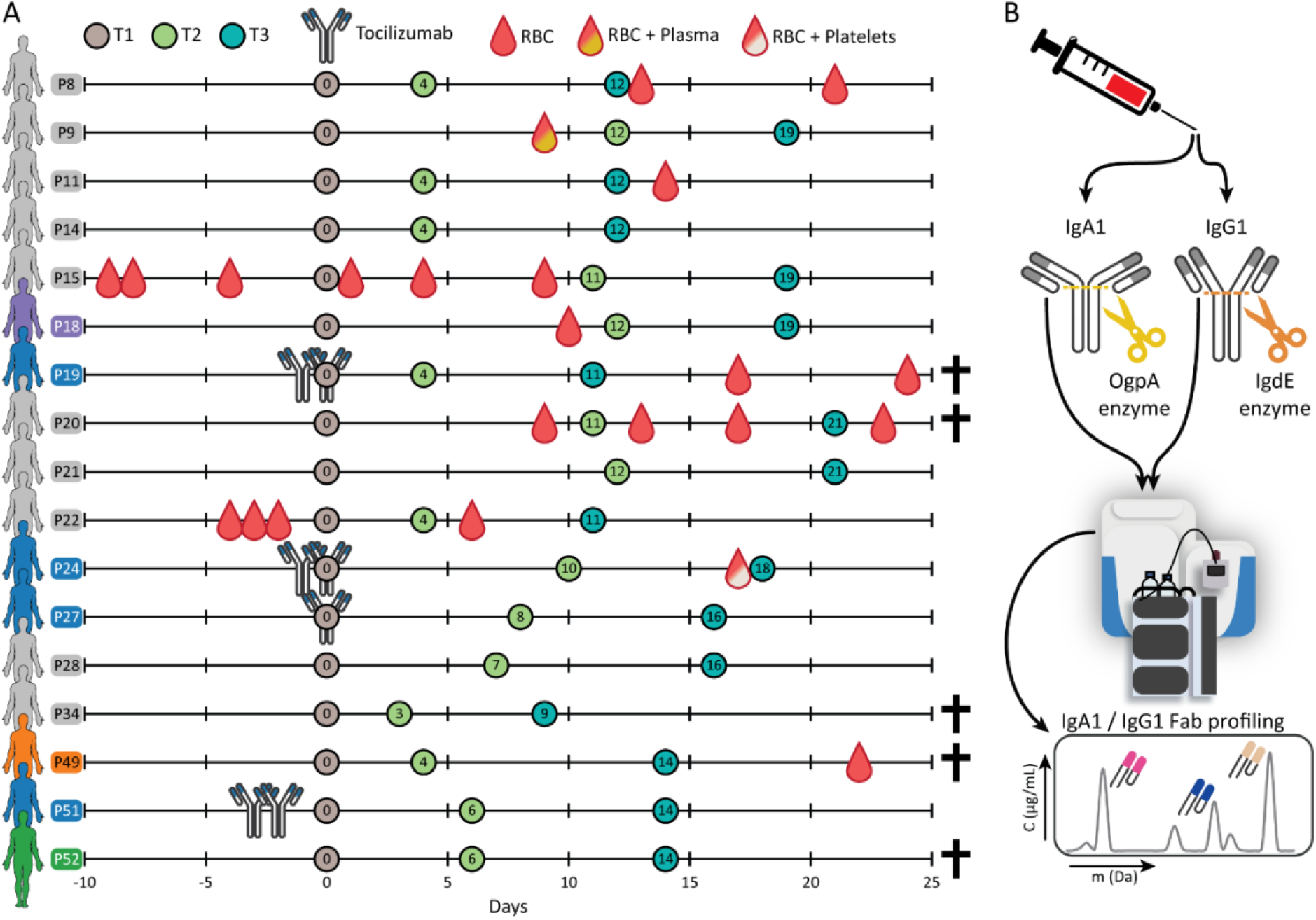
Schematic overview of the cohort, the blood sampling, and the applied method for repertoire profiling. **A)** Timeline of blood sampling with annotated relevant treatments, namely monoclonal antibody therapy (tocilizumab), red blood cell transfusion (RBC), red blood cell and plasma transfusions (RBC + Plasma) and red blood cell and platelet transfusions (RBC + Platelets). For each patient (annotated as Pxx), the first blood sample was marked as day 0 (brown circle; T1). The consecutive sampling time points T2 (green circle) and T3 (blue circle) are annotated with the number of days after T1. Several patients are highlighted in color, these individuals are discussed in more detail: blue when receiving tocilizumab monoclonal antibody therapy, orange when the repertoires were analyzed more in depth, and one donor in green for which we measured a completely aberrant serum profile at T3. Patients that did not survive are annotated with a cross. **B)** Schematic overview of the IgG1 and IgA1 profiling methodology. IgA1 and IgG1 are captured from individual’s blood serum and digested in the hinge region by specific enzymes OgpA and IgdE. The released Fab fragments are then separated and analyzed by intact LC-MS, resulting in either an IgA1 or IgG1 clonal profile.

The cohort was not designed for this study but does reflect a real-life situation in a clinic at the onset of a pandemic. By applying our methods to monitor – longitudinally and in parallel – the IgG1 and IgA1 repertoires of these patients, we aimed to address the question of how antibody repertoires react during changes in physiology of these patients. As these patients all suffered from an infection by the early variant of SARS-CoV-2 we hypothesized to observe the rise and presence of SARS-CoV-2 targeting antibodies in their serum. However, we were equally interested to see whether the unique individual clinical treatment each patient received influenced their serological Ig repertoires over time.

## Results

Using experimental approaches detailed previously we purified either IgG or IgA from the serum of the donor and used subsequently the enzyme IgdE or OgpA, to cleave off and extract the Fab fragments of all IgG1 and IgA1 clones, respectively (Bondt, Dingess et al. 2021, Bondt, Hoek et al. 2021). These Ig Fab fragments cover all six hypervariable CDR regions of the heavy and light chain, which results in each Fab having a characteristic mass in the range of 45-49 kDa. Separation of all these Fab molecules and analyzing them by intact mass LC-MS provides a view at the IgG1 or IgA1 clonal repertoire. By spiking in recombinant mAbs of known quantity, the concentration of each of the detected endogenous Fabs can be obtained. In this manner, the recorded Ig profiles provide a qualitative and quantitative measure of the 50-500 most abundant antibodies in the serum of the donors. Having 17 donors affected by COVID-19, and sampling blood over three time points while being hospitalized, we in were able to record 102 Ig profiles, 51 for IgG1 and 51 for IgA1. As analytical controls we added two (non-related) healthy donors of which we also had samples at 3 time-points (at one-month intervals), extending our analysis to in total 108 Ig profiles.

### Huge variety in total levels of IgG1 and IgA1 in individual patients

Besides information on the concentration of each individual Ig clone in the repertoire our data also provides a means to measure the total levels of IgG1 and IgA1 in the individual patients at each time point of sampling. We quantified the total levels of IgA1 and IgG1 by summing up the total concentrations of Fab molecules derived from the repertoires. The total IgG1 and IgA1 levels in each donor were found to vary widely, ranging from 0.5 to 25 mg/mL for IgG1 and 0.1 to 4.5 mg/mL for IgA1. These IgG1 and IgA1 levels do not necessarily correlate per donor as IgG1/IgA1 ratios also ranged from 0.3 to 25, with, quite remarkably, several donors exhibiting higher IgA1 levels than IgG1 (Figure 2A and Supplemental Table 1). To illustrate these variations, we highlight the levels of a few selected patients in Figure 2A, from which it is apparent that we detected for P51 consistently equal amounts of IgG1 and IgA1 in serum over all time points of around 0.85 +/- 0.2 mg/mL, and this donor thus had roughly 1.7 mg/mL of (IgG1+IgA1) in the serum, and an IgG1:IgA1 ratio of around 1. In contrast, for donor P52 the IgA1 levels were consistently around 4.25 +/- 0.25 mg/mL, whereas the IgG1 levels were roughly 10.5 +/- 1.5 mg/mL. This donor thus had approximately 15 mg/mL of (IgG1+IgA1) in the serum, and an IgG1:IgA1 ratio of around 2.5. Quite peculiar, donor P18 showed IgA1 levels that decreased marginally over time from 2.6 to 1.4 mg/mL, whereas the total IgG1 levels were below the total IgA1 levels at timepoint 1 and 2 (0.86 and 1.48 mg/mL respectively). However, at timepoint 3 the IgG1 concentration had risen to 6.7 mg/mL, 5-fold higher than the total IgA1 concentration at this latest timepoint. By monitoring these total IgG1 and IgA1 levels in the serum of the donors we already observed that patient’s serological Ig levels are widely variable.

**Figure 2.**
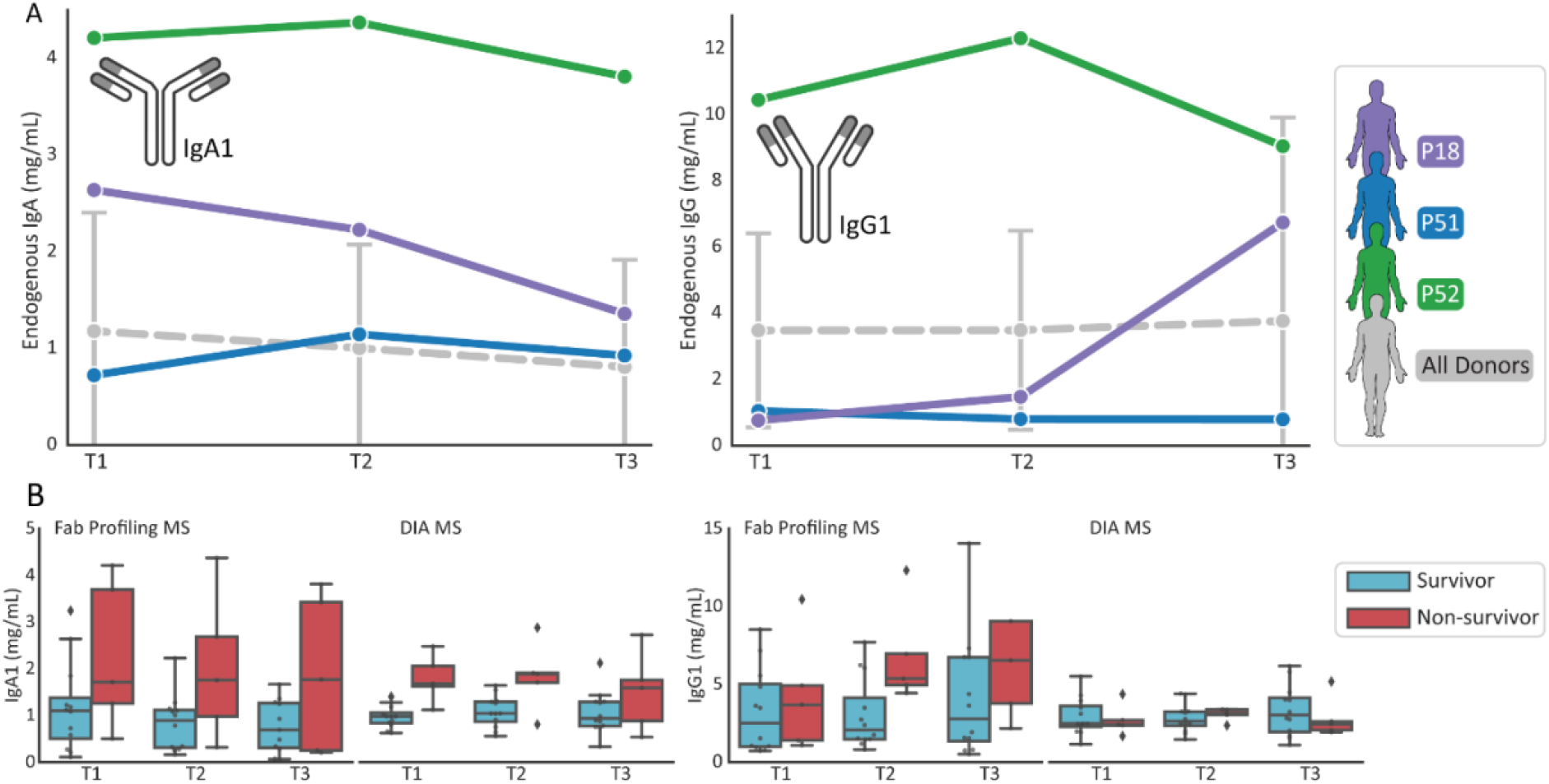
Total levels of IgG1 and IgA1 in individual patients. **A)** For three patients (P18, P51 and P52) levels of IgA1 (left) and IgG1 (right) are depicted as determined by direct Fab profiling LC-MS (FP-MS), illustrating high variability. In grey the average concentration of all donors is plotted. **B)** Comparison of total levels of IgG1 and IgA1 as measured by FP-MS and by shot-gun data-independent acquisition (DIA-) MS. For quantification by FP-MS the sum of all unique Fab concentrations was taken. For shot-gun proteomics quantification was based on unique peptides of the constant regions of either IgG1 or IgA1. Although the data for the FP-MS approach are limited to a sub-cohort of the data gathered for the DIA-NN approach, the results are very much in agreement with each other, indicating that IgA1 levels in non-survivors are on average relatively higher than in survivors, especially at the earliest timepoints. Diamonds outside the box and whiskers indicate outlier values.

Several previous serum proteomics studies using mass spectrometry-based peptide-centric (bottom-up) approaches revealed a decreased survival rate in COVID-19 patients with higher IgA1 levels (Demichev, Tober-Lau et al. 2021, Geyer, Arend et al. 2021, Vollmy, van den Toorn et al. 2021). In **Figure 2B** we present a comparison of our quantification with the data from the study of Völlmy *et al*. who measured IgA1 and IgG1 levels in a larger subset of the same cohort of 17 survivors and 16 non-survivors (Vollmy, van den Toorn et al. 2021). Our current findings corroborate the earlier serum proteomics data, as we here also observe that IgA1 levels in non-survivors are higher at time point 1 and 2 and possibly also at time point 3. Overall, we observe a consistency between the serum proteomics data and Fab-profiling based data on the levels of IgG1 and IgA1, which further validates both the reproducibility in protein quantification by using these two very different methods, and the association of high IgA1 levels with poor COVID-19 outcome, at least in severe hospitalized patients.

### IgA1 and IgG1 clonal profiles are unique for each donor

We next compared all the clonal profiles of IgG1 and IgA1 qualitatively and quantitatively. For each clone with a specific retention time and molecular weight (^[RT]^[Clone#]_[Mw]_) we assessed its concentration and compared it to clones having exactly the same ^[RT]^[Clone#]_[Mw]_ in all the other profiles. Interestingly, but in agreement with what we reported earlier, each unique clone was generally only detected in a single donor, albeit then mostly at all three time points of blood sampling. To provide an overview of all the data we used a correlation score comparing each IgG1 or IgA1 profile. In Figure 3 a correlation matrix is shown comparing on the left all IgA1 profiles and on the right all IgG1 profiles, obtained for all 17 patients (and the two healthy controls) at each of the 3 consecutive time points.

**Figure 3.**
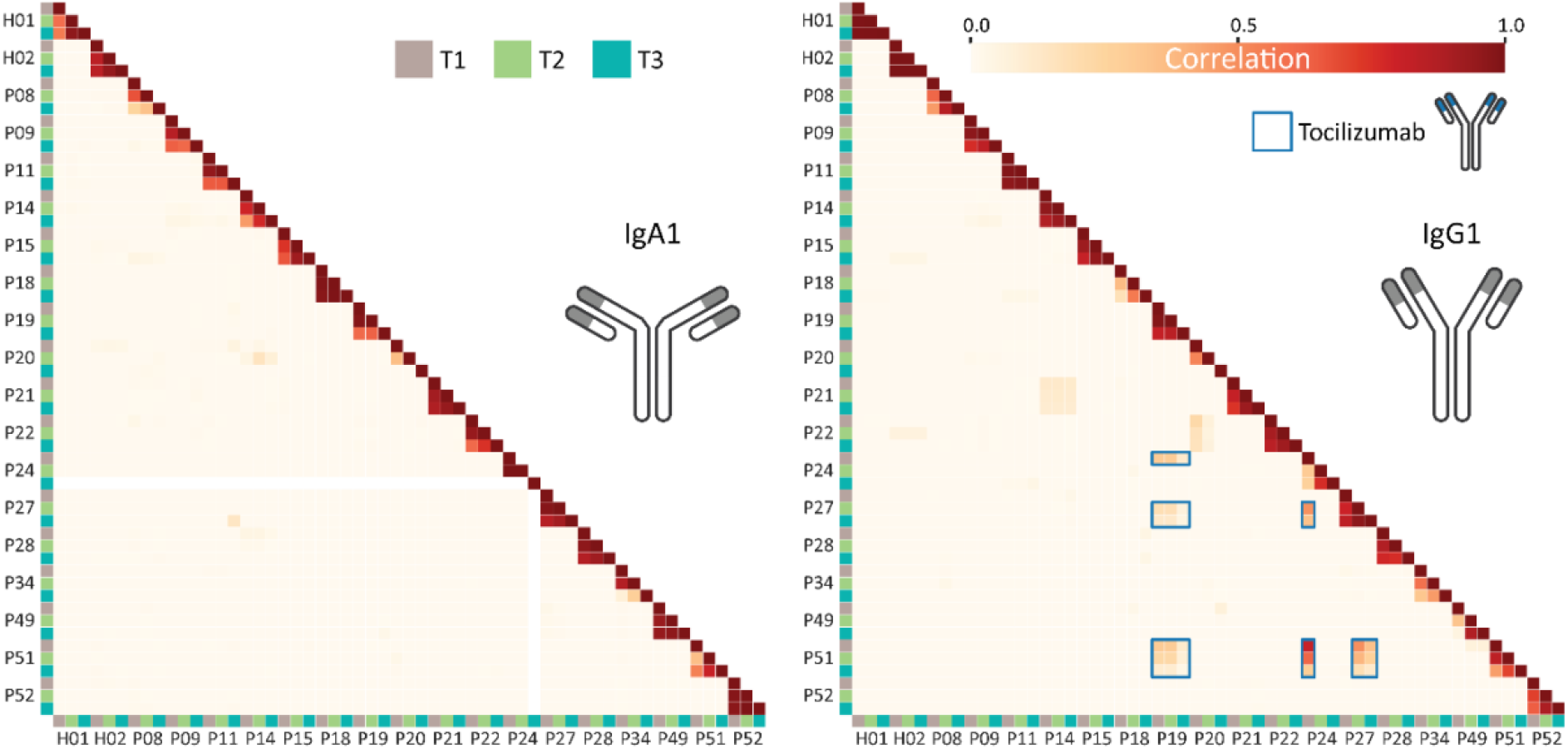
IgA1 and IgG1 clonal profiles are unique for each donor. Correlation between patient antibody repertoires for IgA1 (left) and IgG1 (right) for each time point as indicated by the color-coded squares (T1: brown, T2: green & T3: blue). These correlation plots are based on concentrations of all clones detected, counting the overlap in clones having identical retention time and mass in the LC-MS Fab traces. The degree of correlation is shown in a color scale from white to red, where a value of zero indicates no correlation and a value of 1 indicates the samples are identical. The IgA1 repertoires reveal close to zero correlation between samples of different patients, but between time points within one patient generally a high correlation is observed. The IgG1 repertoires show a similar trend as IgA1, with some notable exceptions indicated by the blue boxes. This unexpected overlap in repertoires originates from high concentrations of a single IgG1 these patients have in common, which we show later to be tocilizumab. Disregarding the tocilizumab IgG1, the data reveal that both IgA1 and IgG1 clonal profiles are explicitly unique for each donor. P24 was included although no IgA data was obtained for T3. At the top H01 and H02 show data obtained from the plasma samples originating from two healthy donors, whose blood was sampled longitudinally at 0, 1 and 2 months, here included as controls.

Nevertheless, during disease development we do observe very substantial changes in the IgG1 or IgA1 repertoires, or both. Clear general trends in these changes within a single patient are not directly observable. For instance, in P52 the IgA1 profiles are nearly identical at all measured time points, whereas the IgG1 profiles show quite dramatic changes over time. In contrast, in P14 the IgA1 profiles do change more over time than the IgG1 profiles. By checking the patients’ medical treatment records, we found that administration of blood products, even including plasma (P9), did not appear to have a significant impact on the Ig repertoires we measured. Comparing for example changes within P8 (no blood products between each of the sampling points) and within P9 (a patient who received RBC + plasma between T1 and T2) there seems to be more changes in the clonal profiles of patient P8 (Figure 3).

### Disease-related drastic changes in the serum IgG1/IgA1 repertoires

Over time several patients started to produce a single or a small number of very abundant IgG1 or IgA1 clones, a feature seemingly more often observed in non-survivors (Figure 4A). As the patients had all recently been infected with SARS-CoV-2 virus, we hypothesized that these prominent changes in the repertoires could originate from antibodies produced to bind and/or neutralize the virus, supported by previous observations that severe ICU patients have higher specific antibody titers (van Rijswijck, Bondt et al. 2022). To probe whether this indeed was the case we used a complementary Luminex bead-based binding assay measuring IgG or IgA binding to a variety of SARS-CoV-2 virus antigens, namely the Spike-trimer, the RBD (receptor binding domain), and the nucleocapsid protein. Of note, these assays probe total IgG or IgA binding, and thus do not distinguish in between IgG1/IgG2/IgG3/IgG4 or IgA1/IgA2. Not surprising, as all patients suffered from severe COVID-19, nearly all patients displayed very strong responses against these antigens (see Supplemental Table 2 and Figure 4B). For many patients these responses were already substantial at T1, but for others a strong response was only observed by T2 or T3.

**Figure 4.**
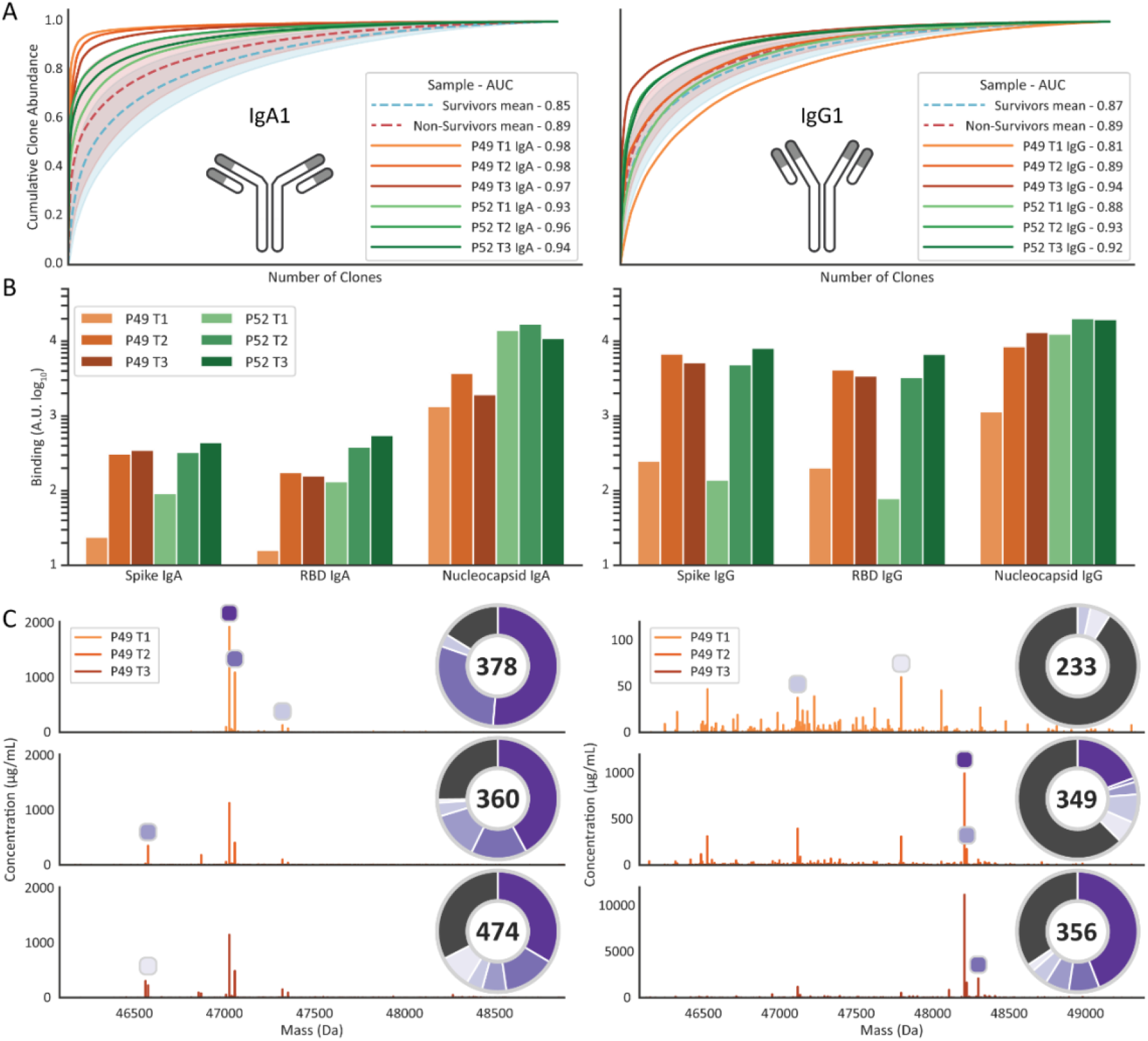
Antibody responses to SARS-CoV-2 can results in clonal profiles dominated by a small number of clones. **A)** Clonal dominance scoring plot. For each serum sample clones are normalized and sorted by abundance. The cumulative abundance of all clones is plotted as curve, and the area under the curve (give in the legend) is a measure of how dominant / dispersed the clonal repertoire of the sample is. The panels show clone dominance scores for IgA1 (left) and IgG1 (right). The red dashed line shows the mean of all non-survivor samples, and the green dashed line for the survivors. The opaque colored areas show one standard deviation from the displayed mean. The two patients with highest clone dominance score have been highlighted, P49 in orange and P52 in green. **B)** SARS-CoV-2 antigen binding monitored by a Luminex assays probing the binding of total IgA (left) or IgG (right) to the antigens; spike-trimer, receptor-binding domain (RBD) and nucleocapsid protein. **C)** Fab mass profiles of P49 IgA1 (left) and IgG1 (right), with relative clone abundances also provided in the pie charts. The pie chart shows the proportion of abundance for each of the top 5 most intense clones, the dark grey slice shows the proportion of the remainder of all other detected clones. The number within the pie chart shows the total number of identified clones within the serum sample. The peaks highlighted in the mass profile correspond with matching color to the slices of the pie chart.

We next focused on P49 and P52 (both non-survivors), as in these patients we observed very abundant IgG1 or IgA1 clones, possibly in response to the virus. In Figure 4B we depict the SARS-CoV-2 antigen binding as observed in the sera of these patients. For the spike and RBD antigen both patients showed a 10- to almost 100-fold stronger response at T2 and T3 compared to T1, and more pronounced for IgG than for IgA. The strong anti-SARS-CoV-2 response should, we hypothesized, also translate in observable changes in the serum IgG1 and IgA1 repertoires of these two patients. In Figure 4C the Fab mass plots for the IgA1 and IgG1 repertoires are shown for P49 and all measured time points. Compared to other mass plots recorded, these plots are remarkable as they are dominated by a low number of clones for IgA1 at all three time-points, whereas for IgG1 the repertoire becomes dominated by only a few clones at T2 and T3 (note the change in y-axis for IgG1). The provided pie-charts reveal that the top 5 clones make up as much as 50 to 80% of all detected Ig molecules in the serum at most time points. In the P49 T3 IgG1 sample, a few clones not present at T1 exhibit serum concentrations of over 1 mg/mL, with the most abundant reaching over 10 mg/mL. The dominant IgA1 clones are already abundant at T1, reaching concentrations of around 1 mg/mL. The observation in this patient that the IgA1 response precedes the IgG response is in line with previous data on SARS-CoV-2 infection (Sterlin, Mathian et al. 2021). The Fab mass plots for the IgA1 and IgG1 repertoires for 52 also display a limited number of dominant clones with concentrations up to 2 mg/mL (Supplemental Figure S1).

### Characterization of the sequences of the putative SARS-CoV-2 targeting IgG1/IgA1s

Through advances in mass spectrometry it has become within reach to sequence recombinant antibodies *de novo* by mass spectrometry using either protein-centric (top-down) or peptide-centric (shotgun or bottom-up) proteomics approaches (Tran, Rahman et al. 2016, Guthals, Gan et al. 2017, Sen, Tang et al. 2017, Peng, Pronker et al. 2021, de Graaf, Hoek et al. 2022, Schulte, Peng et al. 2022), or combinations thereof. However, to do this on endogenous serum clones, in a background of many highly similar antibodies, represents still an arduous undertaking. Particularly, in bottom-up proteomics – whereby proteins are digested into smaller peptides – it remains difficult to connect the peptide sequencing data to the antibody of origin, since large portions of antibody sequences are shared between all clones. Top-down and middle-down approaches targeting the intact Fab or LC and Fd fragments may then come to the rescue, especially as some fragmentation methods, such as electron-capture dissociation (ECD) and electron-transfer dissociation (ETD), produce sequence tags covering the highly variable clone-specific CDR regions (Mao, Valeja et al. 2013, den Boer, Greisch et al. 2020, Shaw, Liu et al. 2020). We recently provided a proof-of-concept that by using an integrative approach combining peptide-centric and protein-centric MS data it is possibly to sequence *de novo* a full (abundant) Fab of a single clone, even in the background of serum (Bondt, Hoek et al. 2021). Here, we followed a similar approach to sequence the most abundant IgG1 and IgA1 clones from the two patients P49 and P52. This approach enabled us to fully sequence the most abundant IgG1 and IgA1 clones as present in the sera of patient P49 and P52 (Figure 5A).

**Figure 5.**
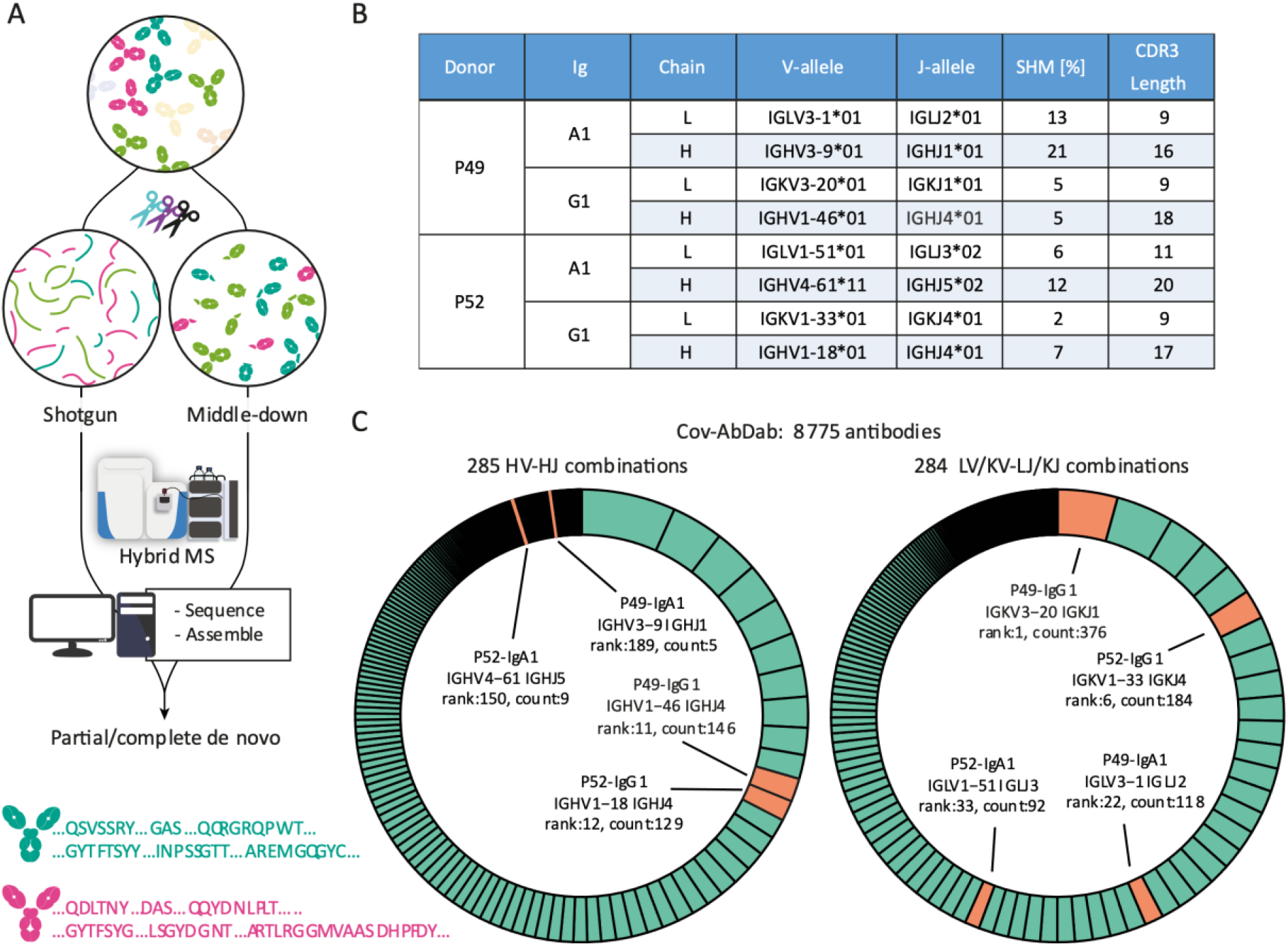
*De novo* sequencing combining peptide- and protein-centric approaches reveal that the dominant clones in P49 and P52 exhibit sequence features alike those deposited in the Cov-AbDab database of antibodies that bind and/or neutralize SARS-CoV-2. **A)** Abundant IgA1 and IgG1 clones were fully *de novo* sequenced using an integrative MS approach, combining peptide-centric (shotgun) and protein-centric (middle-down) approach. In the peptide-centric approach the Fab molecules were digested in parallel with a mix of proteases to provide a set of large set overlapping peptides covering the sequence of the Fab. In the protein-centric MS approach the Fab molecules mass-selected and fragmented with electron-transfer dissociation (ETD) providing sequence tags and mass-restraints that were used to score and refine the sequences predicted form the bottom-up approach. The protein-centric approach was performed both at the intact Fab-level and following reduction of the disulfide bridges in the Fab, by mass-selecting ions of either the Fab light-chain or Fd heavy chains. **B)** Overview of the best matching germline alleles, the amount of somatic hypermutations and the CDR3 lengths for the fully sequenced most dominant IgG1 and IgA1 clones from the serum of P49 and P52. **C)** Assignment (depicted in orange) of the IGHV-IGHJ and IG[K/L]V-IG[K/L]J gene pairs from the here determined clones (see 5B) among the unique pairs in the database of SARS-CoV-2-related antibodies (8775 total entries in Cov-AbDab, with categories depicted in green in the donut charts). The antibodies in the Cov-AbDab database are primarily discovered by B cell sequencing. Especially the germline sequences from the IgG1 clones identified in this study match well within some of the most copious categories in the Cov-AbDab database.

In this case, by using just the protein-centric middle-down approach we obtained confident identification of the closest related germline sequences. This revealed that the light chains of these clones possess limited divergence from the germline sequences. For the more diverged Fd chains, we additionally relied on the peptide-centric data to support or correct the protein-centric germline sequence prediction. Integration of the protein- and peptide-centric data allowed us to obtain accurate sequences, as corroborated by very close (within 1 Da) mass matches (Supplemental Table S3) and extensive coverages of the final sequences (Supplemental Figure S2). Compared to the germline templates, the mature sequences present in the serum of P49 and P52 showed varying degrees of somatic hypermutation, with IgG1 molecules demonstrating less variability than IgA1 molecules (Figure 5B). The CDR3 lengths were as expected longer for the V-D-J-gene encoded Fd chains (16-20 residues) than for the V-J-gene encoded light chains (9-11 residues).

Next we explored how the here derived sequences compared to the 8775 human sequences deposited in the SARS-CoV-specific database of antibodies (Cov-AbDab, version of 26^th^ July 2022; (Raybould, Kovaltsuk et al. 2021)). By looking at the frequencies of unique IGxV and IGxJ combinations, we estimated the gene usages among these 8775 deposited human antibodies. Among the 285 and 284 unique entries (for light and Fd chains, respectively), the here obtained germline sequences for the patient IgG1 molecules scored very well, ranking as top-10 or top-20 for the Fd and light chains, respectively (Figure 5C). When we searched the Cov-AbDab repository for unique light-heavy combinations that match the clones from this study, we found that the best scoring IgG1 clone from P49 again showed a high-ranked match (Supplemental Figure S3). In contrast, the germline sequences here determined for the IgA1 molecules were less frequently present in the Cov-AbDab database.

Although we do not yet have unambiguous prove that the IgG1 and IgA1 clones sequenced here by us have binding or neutralizing activity against SARS-CoV-2 virus, we believe that the Luminex assays data and data from this meta-analysis against the Cov-AbDab database make them very likely candidates.

### Quantitative monitoring of tocilizumab in the serum IgG1 repertoire

The data on IgG1 and IgA1 clonal repertoires revealed that individuals hardly share any Fab clones. Therefore, we were intrigued to detect a substantial overlap between the IgG1 profiles of a small subgroup of patients (namely P19, P24, P27 and P51; Figure 3). Closer inspection revealed that this overlap could be fully attributed to a single clone, whose Fab fragment had in the LC-MS data consistently a retention time of 18.6 min and an Mw of 47441 +/- 1 Da. This single clone was in some of these patients (by far) the most abundant antibody, explaining the high observed overlap in the IgG1 repertoires of these patients, as depicted in Figure 3. We hypothesized that this could be due to treatment of these patients with a therapeutic monoclonal IgG1. Indeed, inspection of the clinical reports for these patients revealed that they had been treated with tocilizumab, a humanized IgG1 acting as interleukin-6 receptor (IL-6R) antagonist. Based on its protein sequence (available on drugbank.com) we found a close match between the mass observed in the patient serum repertoires and the theoretical mass of the tocilizumab Fab (Mw = 47442.02 Da), accounting for the glutamine to be in a pyroglutamate form. Tocilizumab is an immunosuppressive monoclonal antibody normally used for the treatment of rheumatoid arthritis and systemic juvenile idiopathic arthritis, and recently also for COVID-19 (Wang, Fu et al. 2021). In our cohort, four patients did receive tocilizumab via intravenous dosing, whereby some of them received repetitive injections (Figure 1). As tocilizumab is a humanized IgG1 antibody, in our experimental approach it will be co-purified with all other IgG1 clonal antibodies present in the serum. Consequently, we detected and were able to quantify the concentration of tocilizumab in parallel to the concentrations of all endogenous IgG1 clones present in the IgG1 repertoire of these patients.

The total concentrations of IgG1 and the concentration of tocilizumab in the patients P19, P24, P27 and P51 as measured in the serum are shown in Supplemental Table S4. In Figure 6A mass plots depict the measured IgG1 profiles of P24, whereby the peak for tocilizumab “clone” is color-coded. The pie-charts shown as insets in Figure 6A depict the relative contribution of the tocilizumab clone to the full IgG1 repertoire, with alike data for the other patients provided in Supplemental Table S4. Strikingly, tocilizumab is often the most abundant antibody in the sera of these patients, with a concentration of 0.20-0.30 mg/mL after a single dose.

**Figure 6.**
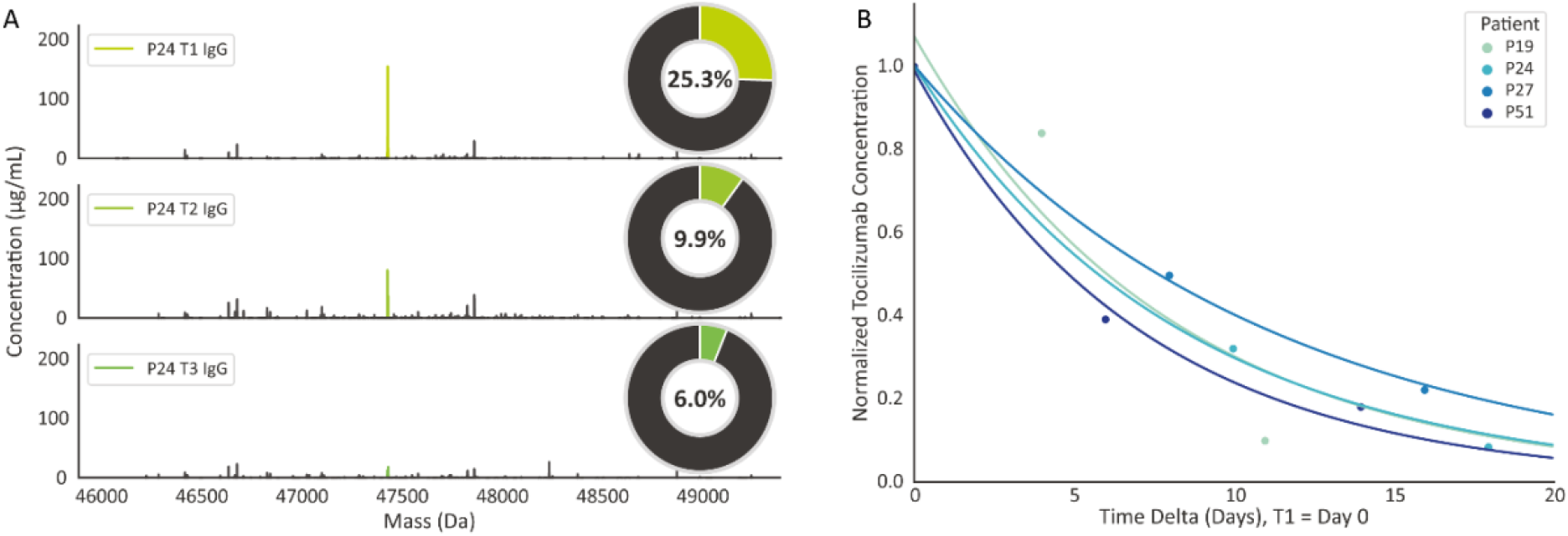
Administered tocilizumab can dominate the IgG1 serum profile but is rapidly cleared. **A)** Deconvoluted Fab mass profiles of P24 at three consecutive sampling points. Each peak represents a unique Fab (based on their unique mass and retention time) and the peak height indicates the clonal concentration. Colored peaks represent tocilizumab, all other peaks are endogenous IgG1 clones. The pie chart shows the contribution of tocilizumab to the total IgG1 repertoire (based on concentrations). **B)** Monitoring Tocilizumab clearance and determination of its half-life in patient serum. T1 was normalized to concentration of 1 and set to day 0. Samples T2 and T3 were normalized to T1. An exponential decay curve was fitted using a non-linear least squares fit. From this curve the half-life was determined to be ∼5.9 days for each of the four patients to which Tocilizumab had been administered.

The FDA recommended intravenous tocilizumab dosing for treatment of COVID-19 is 8 mg/kg tocilizumab with a maximum dose of 800 mg, dependent on disease severity. The patients in our cohort received between 2.4 and 9.4 mg/kg tocilizumab. The total volume of blood in adult human beings can be estimated using the formula 70/sqrt(BMI/22) mL/kg (Lemmens, Bernstein et al. 2006), of which approximately 50 percent is serum. Thus, with a dose of 640 mg, the initial concentration of tocilizumab would be approximately 0.27 mg/mL in a patient with a BMI of 23 (e.g. patient P19), or 0.19 mg/mL in a patient with a BMI of 38 (e.g. patient P24). Among the patients and serum samples we measured the tocilizumab concentrations were in between 0.03 and 0.55 mg/mL (Supplemental Table S4). In all these patients we did observe a decay in abundance of tocilizumab over time, following the intravenous initial dosing. We looked at this decline (Figure 6B), normalizing for each patient on the abundance at T1. Pleasingly, in all these patients we detect a similar decline in tocilizumab abundance, from which we estimated an average half-life of tocilizumab in the serum of these older and severe COVID-19 patients of about 5.9 days. These findings agree well with data from the Centre for Evidence Based Medicine (https://www.cebm.net/covid-19/tocilizumab/).

### Defining an outlier serum sample by IgG1 and IgA1 repertoire profiling

IgG1 and IgA1 repertoires are very unique for each individual, not changing that much over time in healthy donors, but adapting to changes in physiology as demonstrated here for patients suffering from COVID-19. Still, even in the more changing repertoires of severely ill donors, these repertoires correlate over time much more with each other than with those of other donors. Therefore, we were somewhat puzzled by the behavior observed for P20 at T3 (Figure 3 and more in detail Supplemental Figure S4A). Both the IgG1 and IgA1 repertoires at T1 and T2 showed a good correlation with each other. In sharp contrast, the IgG1 and IgA1 repertoires measured for the sample taken at T3 contained no clones that did overlap with the repertoires measured at T1 and T2. Assessing the antibody responses of these sera to the SARS-CoV-2 antigens also revealed that the serum at T3 was a real outlier, as it did not reveal any significant response against any of the probed antigens, whereas these responses were substantial at T1 and T2 (Supplemental Figure S4B). From all this data we need to conclude that the serum annotated as T3 for patient P20 was very likely misannotated and originated from another donor, seemingly not infected with SARS-CoV-2. Although this is evidently an unwanted result, it confirms again clearly that both IgG1 and IgA1 repertoires are highly unique and personalized, and may even be used to track serum samples back to individual patients. Notably, such a miss-annotated serum sample is easily picked up by the current Ig repertoire approach, but harder to elucidate in more standard serum proteomics studies. With this hindsight we did remove this time point from our earlier analysis (Vollmy, van den Toorn et al. 2021), and fortuitously discovered that exclusion of this time-point did not change our major findings. As a general warning, it can be questioned whether such a mix-up of samples is exclusive for our study, or whether other even much higher high-throughput serum proteomics studies may be troubled by such “human mistakes”. Fab profiling, as shown here, can in such cases help as it provides donor unique signatures.

## Discussion and Conclusion

Only recently it has become feasible to obtain detailed insights into the circulating human antibody repertoire. We and others have shown that these repertoires are unique for every donor, and that they may differ from the repertoire as observed from B cell analysis (Williams, Ofek et al. 2017, Bondt, Dingess et al. 2021, Bondt, Hoek et al. 2021). Our previous studies involved patients with a variety of infecting agents, or healthy subjects. Therefore, in the current study, we aimed to rule out that the diverse response to infection was due to the infecting entity by including patients infected by the same pathogen (here: SARS-CoV-2). Again, the donors show very different responses. We see for several donors a strong and early recognition of virus proteins by IgA, whereas a strong IgG1 response occurs somewhat more delayed (Supplemental Table 2), which is in line with previous reporting on the SARS-CoV-2 response (Sterlin, Mathian et al. 2021). However, in a few donors the opposite is observed (i.e. early IgG1 response), or we even do not see an obvious response. The detected IgG1 and IgA1 repertoires change over time, but the amplitude of these responses and their time course seems to be a highly personalized feature. It is therefore most likely that other immunological factors, presumably donor-specific, are regulating personalized responses to a COVID-19 infection. Intriguingly, several studies have previously shown a decreased COVID-19 survival rate in correlation with high IgA1 levels (Demichev, Tober-Lau et al. 2021, Geyer, Arend et al. 2021, Vollmy, van den Toorn et al. 2021), similar as our findings presented in Figure 2B. On the other hand, we present in Figure 4A that the non-survivors score higher in clonal dominance, in other words, they more often have a small number of clones dominating their IgA repertoire. This suggests that potentially only a limited number of clones is present in harmful concentrations or acts in the immune response in a harmful manner, and not that high IgA1 in itself is causing poor survival.

For a few of the dominant IgG1 and IgA1 antibodies we were able to obtain *de novo* sequencing information by a combination of bottom-up and top-down mass spectrometry approaches. When comparing these sequences with those deposited in the Cov-AbDab database, we found that especially the IgG1 clones had underlying germline sequences (IGxV + IGxJ genes) that were among the top-10 and top-20 for the light and heavy chains, respectively, when compared to the database of nearly 10000 human antibody entries (Han, Wang et al. 2021, Raybould, Kovaltsuk et al. 2021). For the here sequenced IgA1 clones, we did not observe an alike SARS-COV-2-specific gene usage. However, the Cov-AbDab database might not provide the same coverage for IgA and IgG isotypes, and sequencing of a higher number of IgA1 clones would be required to draw conclusions on the immunological role of this isotype in SARS-COV-2 infection. Furthermore, the sequences stored in the database are predominantly Spike-protein directed, whereas the IgA response has been shown to be stronger against the nucleocapsid protein than against Spike, particularly in the early phase (Sterlin, Mathian et al. 2021, Kurano, Morita et al. 2022). Summarizing, although factors determining which antibody isotype is used against a certain pathogen may be unknown, when IgG1-based responses are triggered our data supports preferential gene usage.

This study and our previous studies have revealed a dominance of only a limited number of clones in serum IgG1 and IgA1 repertoires. To some this raised the question whether what we see is not just the so-called “tip of the iceberg”. In the current study we therefore compared the Fab-based quantitation with our recent quantitative DIA dataset of the same samples. We found a fairly high similarity in the quantities of IgG1 and IgA1 as determined by the two methods. These two complementary methods of quantitation also revealed that IgG1 and IgA1 concentrations in individuals may deviate substantially from the textbook-reported concentrations. Of note, the current study involves elderly donors, and increased age has been associated with lower antibody repertoire diversity (Tabibian-Keissar, Hazanov et al. 2016). Whether the quantitation matches this well in donors of younger age is yet to be determined.

In addition to the biological hypothesis-generating findings described in this manuscript, we found proof of the value of our method for monoclonal antibody treatment pharmacokinetics. Without prior knowledge we identified a common clone in multiple donors. This turned out to be tocilizumab, a therapeutic monoclonal antibody. Pharmacokinetics of human antibodies are difficult to study because of the high similarity with endogenous antibodies. Therefore, specific detection protocols need to be established for each compound, often including the generation of for example anti-idiotype antibodies (Ovacik and Lin 2018). Using the approach presented here any IgG1 monoclonal antibody – or even combinations thereof – can be detected and quantified over time to determine the relation between dose and serum concentration, as well as *in vivo* half-life like we showed here for tocilizumab. Furthermore, the technology could be used to support therapeutic drug monitoring (TDM) guided clinical decision making (Oude Munnink, Henstra et al. 2016), especially in combination with size exclusion chromatography, as demonstrated in our previous work (Dingess, Hoek et al. 2022), This aids to focus on functionally active mAbs (e.g. not in complex with antigen or anti-drug antibodies), a feature not often possible with other techniques (Ovacik and Lin 2018).

To conclude, we show that donors’ responses against a common pathogen are highly personalized. Also, we provide evidence for the functional application of our methodology in a pharmaceutical or clinical setting for the monitoring of monoclonal antibody therapeutics. The case presented here advocates that clinical diagnostic tools need to be further advanced and become more personalized, as each person has its own response to pathogens and diseases, but also react differently to treatments. We suggest that monitoring antibody repertoires in patients may be one of the more useful clinical tools to monitor serological immune responses in patients due to illness, medical treatment or treatment-resistance.

## Data Availability

All data produced in the present study are available upon reasonable request to the authors

## Acknowledgements

We acknowledge support from the Dutch Research Council (NWO) funding the Netherlands Proteomics Centre through the X-omics Road Map program (project 184.034.019) and the EU Horizon 2020 program INFRAIA project Epic-XS (Project 823839). This research was funded by the Dutch Research Council NWO Gravitation 2013 BOO, Institute for Chemical Immunology (ICI; 024.002.009), and the Utrecht Molecular Immunology Hub.

## Methods

### Patients and Methods Ferrara patient cohort serum sample collection and chemicals

The present analysis included patients from the “Pro-thrombotic status in patients with SARS-CoV-2 infection” (ATTAC-Co) study (Campo, Contoli et al. 2021, Contoli, Papi et al. 2021, Spadaro, Fogagnolo et al. 2021, Vieceli Dalla Sega, Fortini et al. 2021). The ATTAC-Co study is an investigator-initiated, prospective, single-center study recruiting consecutive patients admitted to hospital (University Hospital of Ferrara, Italy) because of COVID-associated acute respiratory distress syndrome between April and May 2020. Inclusion criteria were (i) age >18 yr; (ii) confirmed SARS-CoV-2 infection; (iii) hospitalization for respiratory failure; (iv) need for invasive or non-invasive mechanical ventilation or only oxygen support. Exclusion criteria were prior administration of P2Y12 inhibitor (clopidogrel, ticlopidine, prasugrel, and ticagrelor) or anticoagulant drugs (warfarin or novel oral anticoagulants), known disorder of coagulation or platelet function and/or chronic inflammatory disease. SARS-CoV-2 infection was confirmed by RT-PCR assay (Liaison MDX; Diasorin) from nasopharyngeal swab specimen. Respiratory failure was defined as a PaO2/FiO2 (P/F) ratio ≤200 mmHg. Clinical management was in accordance with current guidelines and specific recommendations for COVID-19 pandemic by Health Authorities and Scientific Societies. Three different blood samplings were made: just after admission (t1), after 7 ± 2 d (t2), and after 14 ± 2 d (t3). Study blood samplings were performed from an antecubital vein using a 21-gauge needle or from central venous line. All patients underwent blood sampling in the early morning, at least 12 h after last administration of anticoagulant drugs. The first two to 4 mL of blood was discarded, and the remaining blood was collected in tubes for serum/plasma collection. The serum and plasma samples were stored at −80°C. The planned blood sample withdrawals were not performed in case of patient’s death or hospital discharge. The ATTAC-Co study population consists of 54 moderate-to-severe COVID-19 patients (Campo, Contoli et al. 2021, Contoli, Papi et al. 2021, Spadaro, Fogagnolo et al. 2021, Vieceli Dalla Sega, Fortini et al. 2021). The subgroup of interest for the present analysis is selected starting from the 16 cases who died and was then narrowed down to 5 patients for which IgG and IgA data was available for all three time points. From the remaining 38 survivors, we identified 17 cases who best matched in terms of age, clinical history, and clinical presentation. This selection was done with the aim of maximizing the possibility to identify differences between deceased and survivors and minimizing potential confounding factors. Of these 17 cases we used data for 12 with all three time points available. The protocol was approved by the corresponding Ethics Committee (Comitato Etico di Area Vasta Emilia Centro, Bologna, Italy). All patients gave their written informed consent. In case of unconsciousness, the informed consent was signed by the next of kin or legal authorized representative. The study is registered at www.clinicaltrials.gov with the identifier NCT04343053.

### IgA sample preparation

IgA Fab samples were prepared as described before (Bondt, Dingess et al. 2021). Briefly, IgAs were affinity captured from patient serum samples using CaptureSelect IgA affinity matrix (Thermo Scientific). Twenty microliter of serum was diluted in 150 µL PBS, and 1 µL PBS containing 200 ng of 7D8-IgA1 monoclonal antibody was added as technical and quantitation reference. After 1 h shaking incubation the unbound fraction of the samples was collected in a fresh tube and stored at 4 °C to be used later for IgG capturing. The bound IgAs were washed and a sialidase cocktail (SialEXO, Genovis, Llund, Sweden) and OgpA enzyme (OpeRATOR, Genovis, Llund, Sweden) were added for overnight digestion of the captured IgA. On the next day, the flowthrough from the digestion containing IgA Fabs was collected after removal of the His-tagged enzymes by Ni-NTA affinity capturing.

### IgG sample preparation

IgG Fab samples were prepared as described before, with minor modifications (Bondt, Hoek et al. 2021). Briefly, IgGs were affinity captured from 85 µL of IgA affinity capturing flowthrough, substituted with 100 µL PBS and 1 µL PBS containing 200 ng trastuzumab and 200 ng alemtuzumab monoclonal IgG antibodies. The monoclonal antibodies were included for retention time and mass reference, and for quantification of all other detected clones. After 1 h shaking incubation the bound IgGs were washed and IgdE enzyme (FabALACTICA, Genovis, Llund, Sweden) was added for overnight digestion of the captured IgG. On the next day, the flowthrough from the digestion containing IgG Fabs was collected after removal of the His-tagged enzyme by Ni-NTA affinity capturing.

### Fab LC-MS(/MS)

Fab LC-MS was performed as described before (Bondt, Hoek et al. 2021), with minor modifications. Briefly, Fabs were separated by reversed-phase liquid chromatography on a Thermo Scientific Vanquish Flex UHPLC instrument with a 1 mm x 150 mm MAbPac RP analytical column. The LC was coupled to an Orbitrap Fusion Lumos Tribrid (Thermo Scientific, San Jose, CA, USA). Separate Fab chains were analyzed with a resolution setting of 120,000 (@ *m/z* 200) in MS1, which allows for more accurate mass detection of smaller proteins (< 30 kDa). MS/MS scans were acquired with a resolution of 120,000. MS1 scans were acquired in a range of *m/z* 500-4,000, and MS2 scans in the range of *m/z* 350-5,000.

### Clonal profiling data analysis

Data analysis of the clonal profile was performed as described before (Bondt, Hoek et al. 2021), with the exception of a few different settings in the BioPharmaFinder deconvolution. Deconvolution was performed using the ReSpect algorithm between 5 and 57 min using 0.1 min sliding window with 25% offset and a merge tolerance of 30 ppm. Using the data of two spiked-in mAbs (trastuzumab and alemtuzumab for IgG1, and 7D8 for IgA1) a mass correction was applied based on the difference between the calculated and observed mAb masses, and similarly, a retention time alignment was applied to minimize deviation between runs. Clones were matched between runs using mass tolerance of 1.5 Da and retention time tolerance of 1.0 min.

### Luminex immunoassays

SARS-CoV-2 specific Luminex Immunoassays were performed as described before (Grobben, van der Straten et al. 2021). Briefly, Spike-trimer, the RBD (receptor binding domain), and the Nucleocapsid protein of SARS-CoV-2 were covalently coupled to Luminex Magplex beads (Luminex) at a ratio of 75 µg protein per 12,5 million beads for Spike-trimer, equimolar quantity for Nucleocapsid protein and 3x equimolar quantity for the RBD. 15 beads per µl were incubated with 1:10,000 diluted serum in blocking buffer (PBS + 2% Bovine Serum Albumin + 3% Fetal Calf Serum + 0.02% Tween-20), overnight at 4°C while shaking. Beads were washed and then stained for 2 hours with either Goat-Anti-Human IgG-PE or Goat-Anti-Human IgA-PE (Southern Biotech). Finally, the beads were washed and analyzed on a Magpix (Luminex). MFI values of bead and buffer only wells were subtracted from the resulting MFI values and the results were presented as arbitrary units (A.U.).

### Bottom-up proteomics for in-depth sequence insights

#### In-gel digestion

Collected Fabs were loaded 3 µg per lane on a 4%-12% Bis-Tris precast gel (Bio-rad) in non-reducing conditions and ran at 120 V in 3-Morpholinopropane-1-sulfonic acid (MOPS) buffer (Bio-rad). Bands were visualized with Imperial Protein Stain (Thermo Fisher Scientific), and the size of the fragments evaluated by running a protein standard ladder (Bio-rad). The Fab bands were cut and reduced by 10 mM TCEP at 37°C, followed by alkylation in 40 mM iodoacetic acid at room temperature in the dark. The Fab bands were digested by trypsin, chymotrypsin, thermolysin and α-Lytic Protease. The digestions were all performed at 37 °C overnight in a 50 mM ammonium bicarbonate buffer. The peptides were extracted with two steps incubation at room temperature in 50% ACN, 0.01% TFA, and 100% ACN respectively. The peptides were dried in speed-vac.

#### Mass Spectrometry

The digested peptides were separated by online reversed phase chromatography on an Dionex UltiMate 3000 (Thermo Fisher Scientific, column packed with Poroshell 120 EC C18; dimensions 50 cm × 75 μm, 2.7 μm, Agilent Technologies) coupled to a Thermo Scientific Orbitrap Fusion mass spectrometer. Samples were eluted over a 90 min gradient from 0 to 35% acetonitrile at a flow rate of 0.3 μL/min. Peptides were analyzed with a resolution setting of 60,000 in MS1. MS1 scans were obtained with a standard automatic gain control (AGC) target, a maximum injection time of 50 ms, and a scan range of *m/z* 350–2,000. The precursors were selected with a 3 Th window and fragmented by stepped high-energy collision dissociation (HCD) as well as electron-transfer high-energy collision dissociation (EThcD). The stepped HCD fragmentation included steps of 25, 35, and 50% normalized collision energies (NCE). EThcD fragmentation was performed with calibrated charge-dependent electron-transfer dissociation (ETD) parameters and 27% NCE supplemental activation. For both fragmentation types, MS2 scans were acquired at a 30,000 resolution, a 4e5 AGC target, a 250 ms maximum injection time, and a scan range of *m/z* 120–2,000.

#### Peptide Sequencing from MS/MS Spectra

MS/MS spectra were used to determine de novo peptide sequences using PEAKS Studio X (version 10.6). We used a tolerance of 20 ppm and 0.02 Da for MS1 and MS2, respectively. Carboxymethylation was set as fixed modification of cysteine and variable modification of peptide N-termini and lysine. Oxidation of methionine and tryptophan and pyroglutamic acid modification of N-terminal glutamic acid and glutamine were set as additional variable modifications. The CSV file containing all the de novo sequenced peptide was exported for further analysis.

#### Template-based assembly via STITCH

The CSV files exported from PEAKS were used as input dataset for STITCH (1.1.2-windows) template-based assembly (Schulte, Peng et al. 2022). The human antibody database from IMGT was used as template. The cutoff score for the de novo sequenced peptide was set as 90 and the cutoff score for the template matching was set as 10. All the peptides supporting the sequences were examined manually.

