## Supplemental Figures for "No patient is the same; lessons learned from antibody repertoire profiling in hospitalized severe COVID-19 patients"

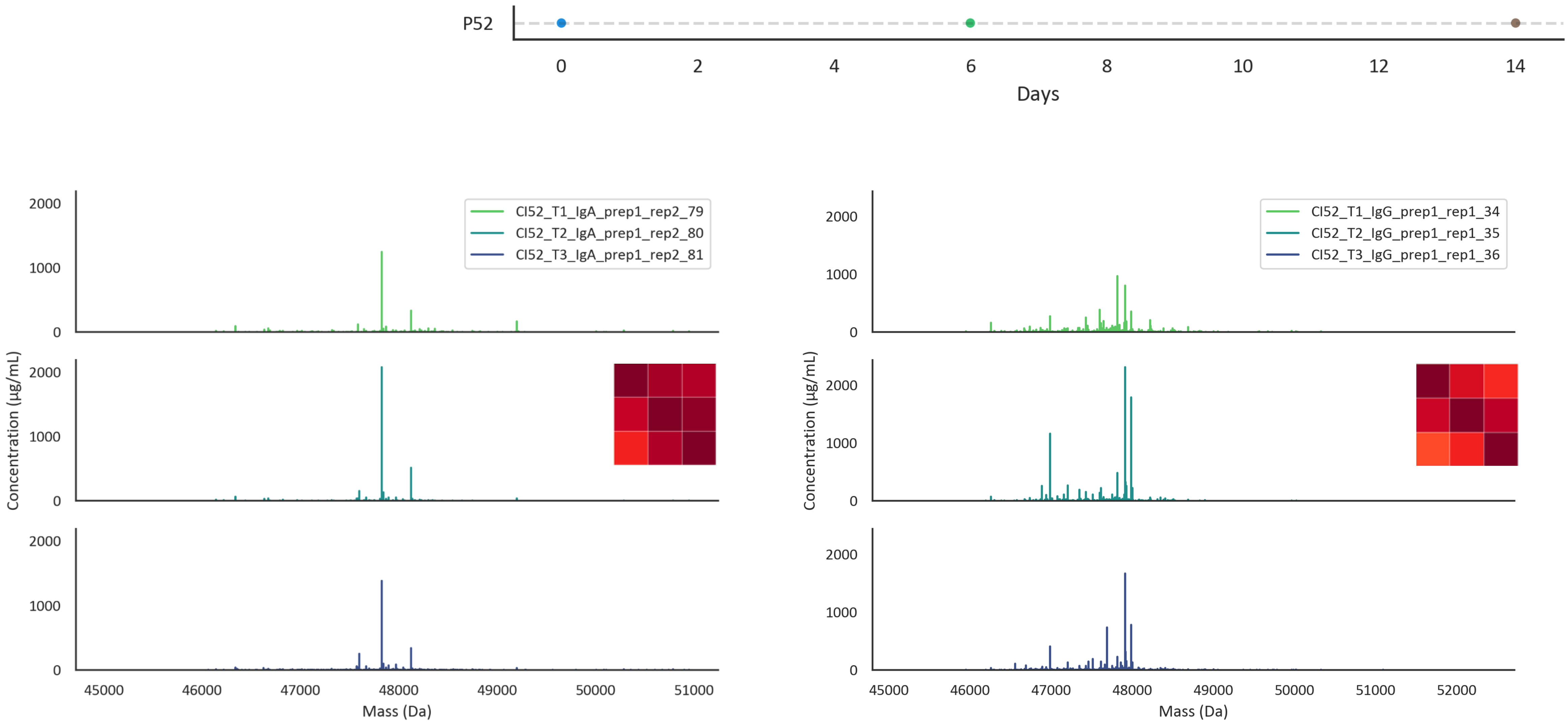


**Supplemental Figure S1 |** Fab mass profiles of P52 showing a limited number of dominant clones, reaching more than 1 mg/mL per individual clone.

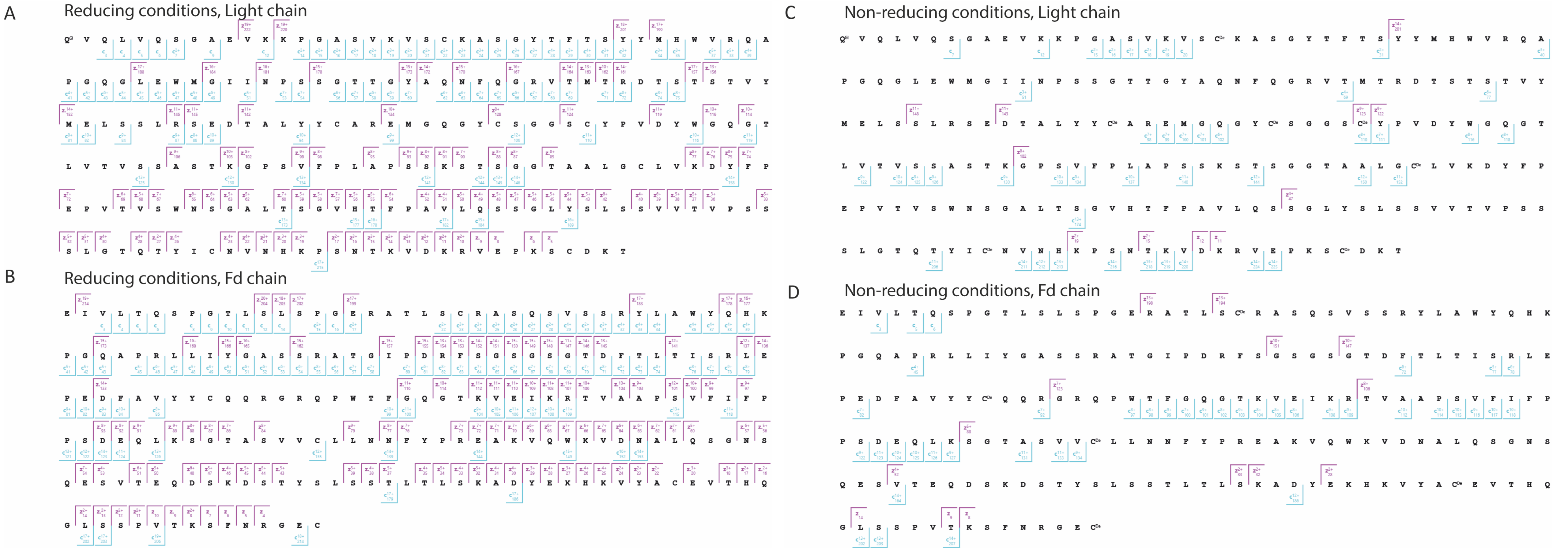


**Supplemental Figure S2 |** Fragmentation maps of the most abundant IgG1 clone from P49 for the light chain (A) and Fd chain (B) after reduction of the disulfide bridges (A, B) and when fragmented as part of Fab (C, D). In all cases MS2 ETD spectra of interest were summed using LcMsSpectator ([https://github.com/PNNL-Comp-Mass-Spec/LCMS-Spectator](https://github.com/PNNL-Comp-Mass-Spec/LCMS-Spectator)) for each condition and c-/z-type fragments were matched using 10 ppm mass tolerance.

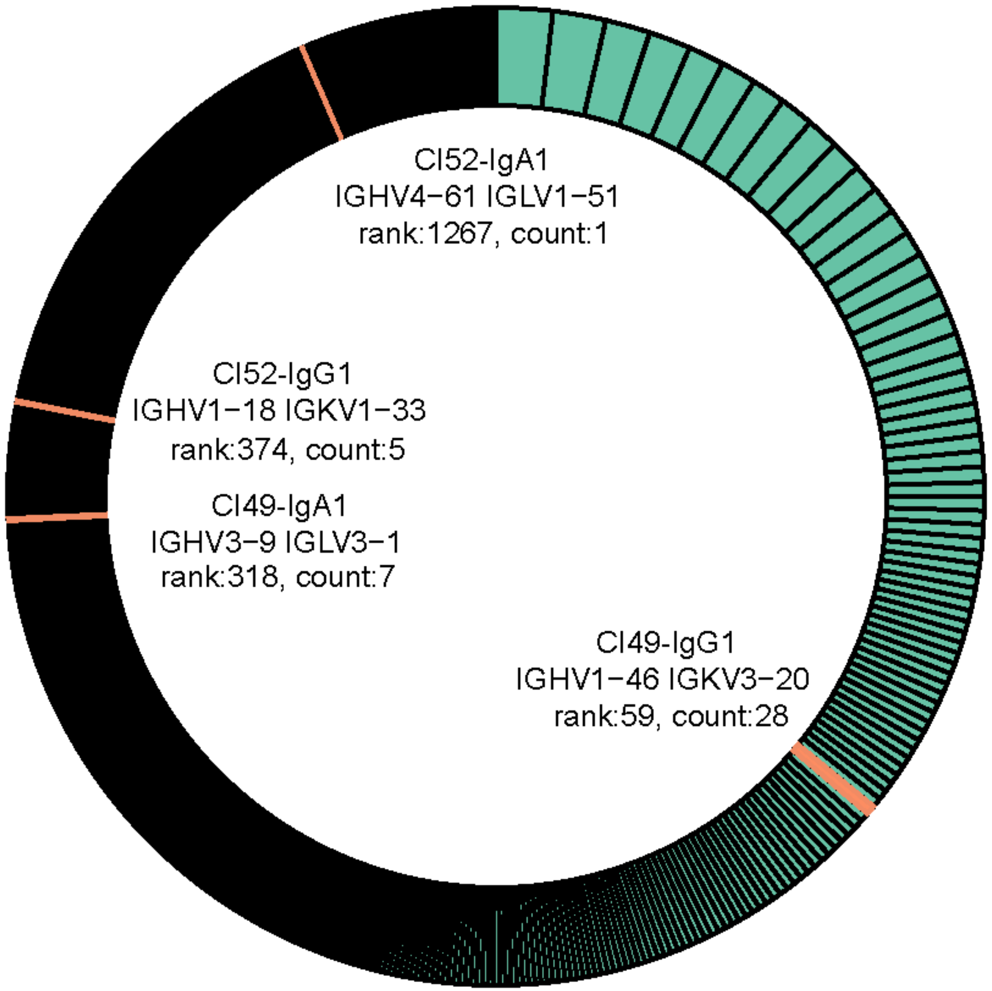


**Supplemental Figure S3 |** Matching of HV-LV combinations to the Cov-AbDab.

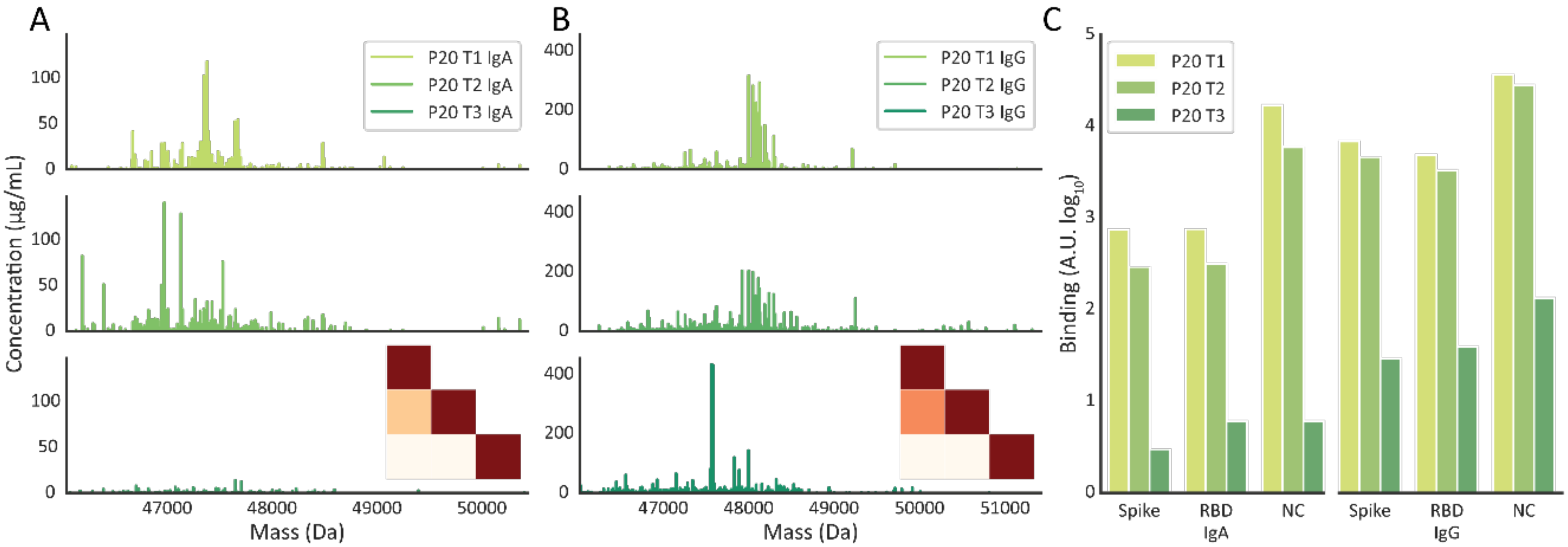


**Supplemental Figure S4 | Defining an outlier serum sample by IgG1 and IgA1 repertoire profiling A**) IgG1 and IgA1 Fab mass profiles of patient P20. Particularly between T2 and T3 the Fab mass profiles of both IgA1 and IgG1 change dramatically, with no clonal overlap at all, resulting in an extremely low correlation. **B**) SARS-CoV-2 antigen binding monitored by a Luminex assays probing the binding of total IgG or IgA to the antigens; spike-trimer, receptor-binding domain and nucleocapsid protein for P20.
